# Spatial Multi-Omics Connect SHROOM3 and COL18A1 in Chronic Kidney Disease

**DOI:** 10.1101/2024.02.09.24301246

**Authors:** Pukhraj S. Gaheer, Nikhil Uppal, Amy Paul, Pedrum Mohammadi-Shemirani, Nicolas Perrot, Guillaume Paré, Darren Bridgewater, Matthew B. Lanktree

## Abstract

Shroom Family Member 3 (*SHROOM3)* encodes an actin-binding protein that impacts kidney development. Genome-wide association studies (GWAS) identified CKD-associated common variants around *SHROOM3* and *Shroom3* knock-out mice develop glomerular abnormalities. We sought to evaluate the impact of genetically predicted *SHROOM3* expression on kidney traits and the circulating proteome, and validate findings in a mouse model. Genetic instruments for *SHROOM3* expression in distinct kidney compartments (glomerular n=240, tubulointerstitial n=311) were constructed using single cell sequencing data from NephQTL2. Using two-sample Mendelian randomization, we evaluated the effects of glomerular and tubulointerstitial *SHROOM3* expression on kidney traits and the concentration of 1,463 plasma proteins in the UK Biobank and CKDGen Consortium. Genetically predicted tubulointerstitial *SHROOM3* expression colocalized with the genetic signals for eGFR and albuminuria. A 34% reduction in genetically predicted tubulointerstitial *SHROOM3* expression was associated with a 0.3% increase in cross-sectional eGFR (*P* = 6.8×10^-4^), a 1.5% increase in albuminuria (*P* = 0.01), and a 2.2% reduction in plasma COL18A1 concentration (*P* = 1.2×10^-5^). In contrast, genetically predicted glomerular *SHROOM3* expression showed neither colocalization nor significant Mendelian randomization results. Using immunofluorescence, heterozygous *Shroom3* knockout mice had a concordant reduction of Col18a1 in their kidneys, primarily around the tubules. Thus, reduced tubulointerstitial *SHROOM3* expression, but not glomerular, is associated with increased cross-sectional eGFR, increased uACR, and reduced plasma COL18A1 and *Shroom3* knockout leads to reduced kidney Col18a1, agnostically linking *SHROOM3* and *COL18A1* in CKD pathogenesis.

**Lay Summary:** The *SHROOM3* gene is consistently linked to kidney disease, but we have yet to fully understand why. We looked at how genetic changes affecting *SHROOM3* impact different regions of the kidney. We found that less *SHROOM3* in kidney tubules led to increased kidney filtration, but also increased leakage of protein into the urine. After looking at over 1000 proteins, we identified a new link between *SHROOM3* and a collagen protein called COL18A1. We then confirmed the link between SHROOM3 and COL18A1 by imaging the kidneys of mice designed to have less SHROOM3. Our results suggest an interaction of SHROOM3 and COL18A1 leads to increased pressure on the kidney filtration system.

## Introduction

Chronic kidney disease (CKD) impacts 10% of the global population, culminating in heightened risk of kidney failure, cardiovascular disease, and death.^1^ Population-scale genome-wide associations studies (GWAS) have identified genetic variants within and around Shroom Family Member 3 (*SHROOM3)* to be associated with CKD, cross-sectional estimated glomerular filtration rate (eGFR), urinary albumin-to-creatinine ratio (uACR), and blood urea nitrogen (BUN).^2-5^ The associated variants are located in a linkage disequilibrium block around the transcription start site of *SHROOM3*, suggesting altered quantity of *SHROOM3* gene expression could play a causal pathophysiological role.^6^ Transcriptome-wide association analyses, exploring links between gene expression and kidney phenotypes, have demonstrated an association between kidney *SHROOM3* expression and cross-sectional eGFR.^7^ Analysis of RNA-sequencing from 50 human kidney transplant biopsies revealed a weak association between *SHROOM3* expression in the glomerular and non-glomerular regions, but exhibiting opposing correlations with eGFR in each region (increased *SHROOM3* expression in glomerular tissue and increased eGFR, *P* = 0.03; decreased *SHROOM3* expression in non-glomerular tissue and increased eGFR, *P* = 0.05).^8^ During embryonic development, both *Shroom3* homozygous and heterozygous knockout mice exhibit abnormal nephron development and a *Shroom3* dose-dependent reduction in glomerular number.^9^ Following acute kidney injury, *Shroom3* heterozygous knockout mice displayed increased mortality, worsened kidney function, and heightened fibrosis.^10^ Studies in different animal models show similar results: post-natal *Shroom3* knockdown induces albuminuria in mice, reintroducing the normal *Shroom3* gene into hypertensive rats alleviates albuminuria and glomerulosclerosis, and introducing wild-type *Shroom3* into *Shroom3*-deficient zebrafish remedies glomerular abnormalities.^11,12^ Despite these insights, the comprehensive impact of the *SHROOM3* gene on kidney disease, relevant biomarkers, and underlying mechanism remains incompletely understood.

Mendelian randomization (MR) is a powerful tool for evaluating causal relationships by utilizing the genetic determinants of a risk factor.^13,14^ If a genetic variant is associated with a lifelong increase in an exposure, and greater exposure is causative for a phenotype, then the variant associated with greater exposure should also be associated with the phenotype. MR has been commonly used to assess traditional risk factors such as lipids, uric acid, hypertension, and body mass index,^15-18^ but can also be used to assess molecular phenotypes such as gene expression as in transcriptome-wide association studies (TWAS), and protein biomarkers as in proteome-wide MR studies (also referred to as proteo-genomic or proteome-wide association studies (PWAS)).^19,20^ Phenome-wide MR further explores the effects of genetically influenced changes in a risk factor on a wide range of phenotypes. By simultaneously investigating a broad range of phenotypes, pleiotropic effects (when a gene or genetic variant influences multiple seemingly unrelated traits) can be investigated to identify shared biological pathways. Colocalization provides evidence for the presence of a shared causal variant between traits, suggesting a shared biological mechanism underlying the association.^21^ MR examines the impact of an expected change in the *quantity* of the risk factor on a specific phenotype. Conversely, rare exonic variant analyses can explore the impact of changes in protein *function*.

We sought to further evaluate the impact of changes in *SHROOM3* quantity and function using population-scale biobank genotyping, sequencing, phenome-wide, and proteome-wide data, coupled with kidney compartment-specific gene expression data, MR, colocalization, and rare variant analyses, with validation of findings in a *Shroom3* knockout model (Figure 1).

**Figure 1:**
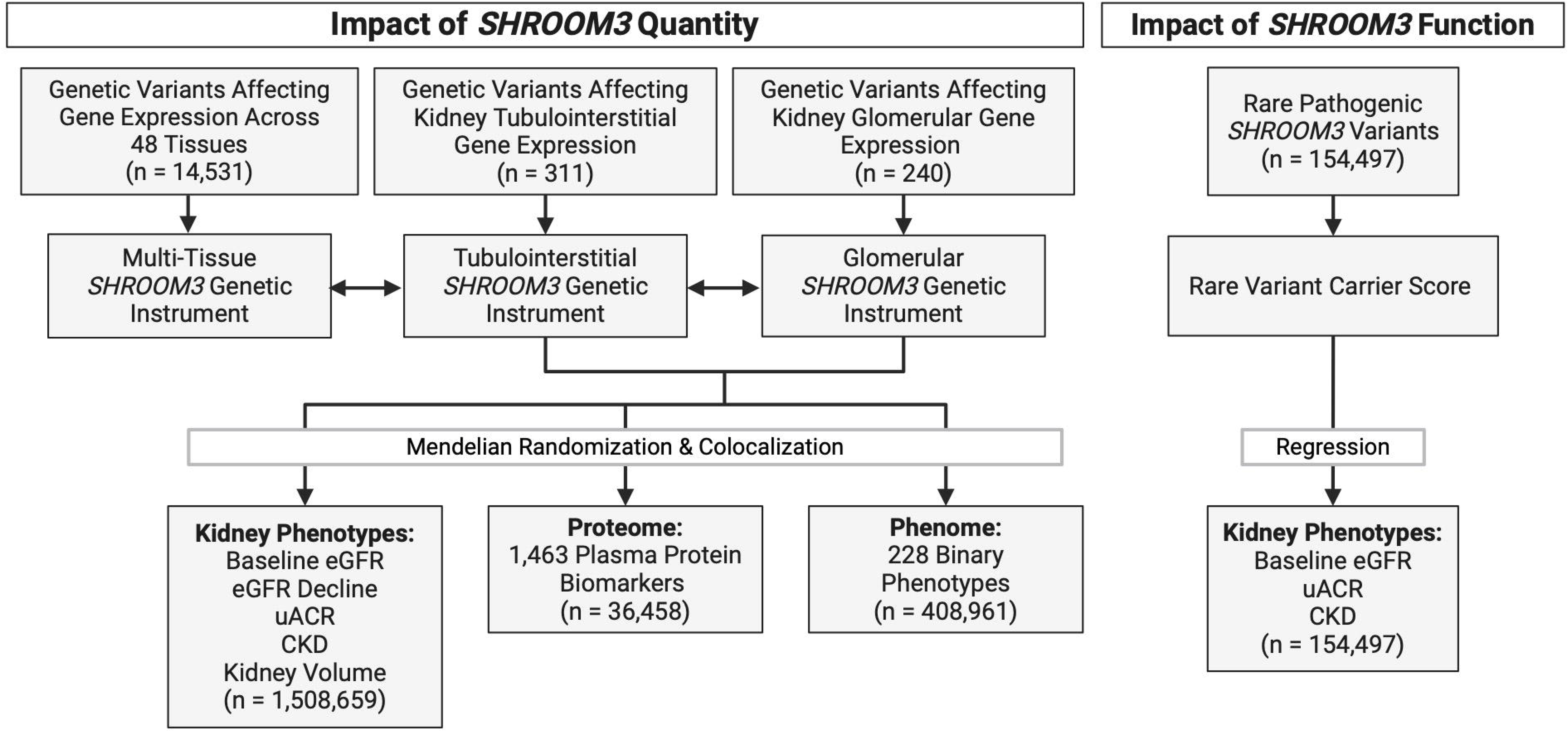
Overall study design of a spatial multi-omic investigation of *SHROOM3* quantity and function. Genetic variants affecting glomerular and tubulointerstitial gene expression were downloaded from GTEx and NephQTL2 database. Kidney phenotype summary statistics retrieved from the CKD Genetics Consortium or calculated in UK Biobank. n, sample size; CKD, chronic kidney disease; eGFR, estimated glomerular filtration rate; uACR, urinary albumin-to-creatinine ratio; BUN, blood urea nitrogen. Created with BioRender.com.

## Methods

### Phenotype Definition in the UK Biobank

All UK biobank outcomes were defined using either: 1) diagnosis of acute, chronic, unspecified, or end-stage kidney disease using the International Classification of Diseases, Tenth Revision (ICD-10) (UK Biobank data field 41270) or algorithmically-defined end-stage kidney disease (UK Biobank data field 42046), or 2) eGFR_Crea_ or eGFR_Cys_ less than 60 ml/min/1.73m^2^ at baseline. eGFR_Crea_, eGFR_Cys_, and eGFR_CreaCys_ (eGFR calculated from serum cystatin C and creatinine) using the 2021 CKD-Epi equations,^22^ or 3) uACR. Urinary albumin was determined in a spot urine sample. uACR and eGFR were log-transformed and standardized. Individual-level analysis was restricted to white British ancestry participants in the UK Biobank. Plasma protein concentrations for 1463 proteins in 36,458 UK Biobank participants were standardized. Data analyses in UK Biobank were performed under application number 15255 and has approval from the Northwest Multi-Center Research Ethics Committee.

### SHROOM3 eQTL Instrument Selection

As *SHROOM3* isn’t expressed in lymphocytes, results from the expression quantitative trait loci (eQTL) genetics consortium (eQTLGen) arising from whole blood were not applicable.^23^ Thus, our first genetic instrument representing multi-tissue *SHROOM3* expression was generated through an inverse variance weighted meta-analysis of eQTLs across 48 tissues in the Genotype-Tissue Expression (GTEx) Project v8. Tissues from GTEx with a sample size of ≥ 70 were included, excluding whole blood due to the low *SHROOM3* expression. This instrument represented genetically predicted *SHROOM3* expression in every tissue.

We analyzed eQTLs from the distinct kidney tubulointerstitial and glomerular compartments using single cell sequencing data from NephQTL2. The Nephrotic Syndrome Study Network (NEPTUNE) cohort is an observational study that enrolls individuals with non-inflammatory glomerular diseases.^24^ Using glomerular and tubulointerstitial data of 332 participants of the NEPTUNE cohort, the NephQTL group identified eQTL genetic variants associated with the expression of 24,474 genes from 240 glomerulus and 311 tubulointerstitial tissue samples.^24^ 55% of participants in NEPTUNE self-reported race as White and 61% of participants were males. We excluded genetic variants more than 500 kilobases away from *SHROOM3*, focusing on cis-eQTLs.^25^ Palindromic and ambiguous variants, and non-synonymous and splicing variants were removed using ANNOVAR and RefSeq annotation.^26,27^ Pruning and thresholding was done within 250 kilobase regions using a linkage disequilibrium threshold of r^2^ < 0.01 to ensure variants were independent. r^2^ was calculated using the 1000 genomes phase 3 European ancestry reference panel.^28^ F-statistic was used to evaluate the strength of each genetic instrument.^29^ Each genetic instrument was passed through Phenoscanner to assess MR assumptions.^30^

### Two-Sample MR

Three assumptions must be met for a MR analysis to be valid: (1) genetic variants should explain a significant proportion of variation in the exposure (relevance); (2) genetic variants in the instrument should be independent of confounders (independence); and (3) genetic variants must not have an alternative direct association with the outcome (exclusion, or absence of pleiotropy).^13^ Careful selection of variants to be included in a genetic instrument and detailed sensitivity analyses are required to evaluate these assumptions. Independent genetic variants not in linkage disequilibrium are combined into scores at the individual-level (individual-level allele score MR), or the effects of numerous independent variants are jointly assessed using regression (summary-level MR).

After generating genetic instruments, outlier variants were removed using MR Pleiotropy RESidual Sum and Outlier (MR-PRESSO) analysis.^31^ Inverse variance weighted (IVW) MR was the primary analysis to infer the relationship between *SHROOM3* expression and kidney traits, with weighted median, MR-Egger, and MR-PRESSO serving as sensitivity analyses.^32,33^ A *P* value < 0.05 for the MR-Egger y-intercept indicates directional pleiotropy. Statistical analyses were performed using R software version 4.3.0 (https://www.r-project.org) with the *TwoSampleMR* package (version 0.5.6).^34^ All analyses were conducted in accordance with STROBE-MR: guidelines for strengthening the reporting of MR studies (Supplementary Table S1).

### Summary-level MR Data

To evaluate eGFR and BUN, we utilized summary-level data from the 2021 Stanzick *et al.* GWAS of cross-sectional eGFR_Crea_, eGFR_Cys_, and BUN (n = 1,201,930) and summary-level GWAS data from 2022 Liu *et al.* trans-ethnic GWAS of eGFR_Crea_ (n = 1,508,659).^3,4^ Diabetes-stratified kidney function was also investigated (n with diabetes mellitus = 178,691).^35^ To evaluate longitudinal eGFR decline we analyzed summary-level data from the 2022 Gorski *et al.* GWAS of longitudinal eGFR decline (n = 343,339).^36^ The eGFR decline GWAS was unadjusted for baseline eGFR, avoiding spurious associations due to collider bias.^37^ Summary statistics from the 2019 Teumer *et al.* European ancestry GWAS of uACR (n = 348,954) and 2021 Liu *et al.*, European ancestry GWAS of kidney volume (n = 32,860) were also analyzed.^5,38^ CKD summary-level data was obtained from the 2019 Wuttke *et al.* CKD Genetics (CKDGen) Consortium GWAS of stage 3 CKD or higher (eGFR_Crea_ < 60 ml/min/1.73m^2^) (41,395 cases and 439,303 controls) which does not include UK Biobank samples.^2^ Thus, we also conducted an inverse variance-weighted meta-analysis of our individual-level MR results with Wuttke *et al*. results.

### Individual-level Allele Score MR

The cumulative effect of these genetic variants was evaluated on 333,735 participants of the UK Biobank by performing linear and logistic regression between individual-level *SHROOM3* genetic risk score and kidney phenotypes adjusted for age, sex, assessment center, and the first 20 genetic principal components of ancestry. Individual-level genotype data was used to create an effect-weighted genetic risk score representing genetically predicted *SHROOM3* expression. For each variant in the genetic instrument, expression-increasing alleles were coded as 0, 1, and 2 and multiplied by the normalized effect on *SHROOM3* expression.

1,463 protein biomarker concentrations were measured in plasma using proximity extension assays developed by Olink (https://www.olink.com; Uppsala, Sweden) in 36,458 European ancestry participants of the UK Biobank. We tested if a genetic risk score representing genetically altered *SHROOM3* expression was associated with altered biomarker concentration, adjusted for age, sex and genetic principal components of ancestry, using two-sample MR analysis. A Bonferroni-adjusted significance threshold for the proteome analysis was 3×10^-5^ (P < 0.05/1463).

### Phenome-Wide MR

To evaluate the breadth of phenotypes impacted by modified *SHROOM3* expression, we performed an MR of 228 binary phenotypes in the UK Biobank. Data was obtained from the 2018 Zhou et al. GWAS catalog.^39^ Analysis included European ancestry participants, phenotypes with a minimal case-to-control ratio of 0.01 and used a Bonferroni-corrected p-value threshold of 2×10^-4^ (P < 0.05/228).

### Colocalization Analysis

Colocalization analysis to examine the overlap of genetic signals for *SHROOM3* expression with eGFR_Crea_, eGFR_Cys_, CKD, uACR, and BUN was performed using the Bayesian colocalization *coloc* R package (version 5.1).^40^ Five hypotheses are tested: null hypothesis (H_0_), no association with either outcome or exposure; H_1_, association with outcome, but not exposure; H_2_, association with exposure but not outcome; H_3_, association with exposure and outcome with distinct causal variants; H_4_, shared causal variants associated with outcome and exposure.^40^ A posterior probability for H_4_ > 0.8 is strong evidence of colocalization. The prior probabilities p_1_ = p_2_ = 10^-4^ and p_12_ = 10^-5^ were set at a conservative threshold.^41^

### Evaluation of Rare Exonic SHROOM3 Genetic Variants

We evaluated the prevalence of rare (minor allele frequency below 1%) predicted loss-of-function (nonsense, frameshift, and splice donor or acceptor) variants by performing a weight-based meta-analysis of allele counts across gnomAD v2.1.1 (non-TOPMED) (122,431 genomes; 13,300 exomes) and BRAVO (132,345 genomes) (bravo.sph.umich.edu/freeze8/hg38/).^42^ An ancestry-specific prevalence estimate of loss-of-function variants was also calculated in gnomAD. Next, rare pathogenic *SHROOM3* variants were tested for association with kidney phenotypes in 157,528 UK Biobank participants with available phenotype and exome sequencing data. Pathogenic *SHROOM3* variants included predicted loss-of-function variants and non-synonymous variants predicted deleterious by the Mendelian Clinically Applicable Pathogenicity algorithm (M-CAP score > 0.025).^43^ The cohort allelic sum test is a rare variant association burden test that aggregates rare genetic variants in an individual into a dichotomous score.^44^ A dichotomous score was incorporated in a linear regression model to test the association between *SHROOM3* rare variant carrier status, and several kidney phenotypes adjusted for age, sex, assessment center, chip type, and the first 20 genetic principal components of ancestry.

### Immunofluorescence Staining of Col18a1

We performed immunofluorescence staining of Col18a1 in *Shroom3* heterozygous knockout mice (*Shroom3*^Gt/+^) previously generated using a gene trap cassette.^45,46^ Whole kidneys from postnatal 3-month-old *Shroom3*^Gt/+^ and CD1 *wild type* mice were resected, fixed in 4% paraformaldehyde at 4^°^C for 48 hours, and embedded in paraffin. Kidneys were sectioned to 5 µm on a Leica microtome, mounted on Superfrost plus microscope slides (VWR, Mississauga, Ontario) and incubated at 37°C overnight. For immunofluorescence, samples were blocked with 7.5% normal goat serum and 4.5% bovine serum albumin at room temperature for 1 hour, then incubated with primary antibody Col18a1 (1:500), overnight at 4^°^C.^47,48^ An incubation buffer containing 1% PBS, 3% bovine serum albumin, 5% normal goat serum, and 0.3% Tween20 was used to dilute the primary antibody. Tissue samples were washed 3 times in PBS, followed by incubation with secondary antibody Alexa Fluor 488 anti-rabbit (Life Technologies, 1:300, Milton, Ontario) in incubation buffer at room temperature for 1 hour, washed in PBS, counterstained with DAPI (Sigma Aldrich, Oakville, Ontario), washed in PBS again, then fixed using Fluoromount mounting medium (Sigma Aldrich, Oakville, Ontario). Images were captured using the Olympus BX80 light and fluorescence microscope, and CellSens image acquisition software.

## Results

### SHROOM3 eQTL Genetic Instruments

Our multi-tissue *SHROOM3* genetic instrument did not colocalize with any of the assessed kidney phenotypes, displaying significant heterogeneity. Thus, we redirected our focus to kidney compartment-specific genetic instruments. After clumping, removing palindromic variants, and removing the variant rs4859682 for violating the exclusion assumption, 12 independent variants were associated with tubulointerstitial *SHROOM3* expression and 10 independent variants were associated with glomerular *SHROOM3* expression (Supplementary Table S2 & S3). For the only overlapping variant, the minor allele (T) of variant rs28584831 reduced *SHROOM3* expression in both kidney compartments and was present in both genetic instruments. Using MR, we observed no association (*P* > 0.7) between either genetic instrument with *SHROOM3* expression in the opposite kidney compartment (Supplementary Table S4 and S8). Both genetic instruments were strong: the tubulointerstitial genetic instrument explained 27% of the variation in *SHROOM3* expression in the kidney tubulointerstitium, and the kidney glomeruli genetic instrument explained 30% of the variation in *SHROOM3* expression in the kidney glomeruli (Supplementary Table S4 & S8).

### Tubulointerstitial SHROOM3 MR Analysis

We found the tubulointerstitial *SHROOM3* expression genetic instrument was associated with cross-sectional eGFR_Crea_ (β = 0.3% increase per 34% relative decrease in *SHROOM3* genetic instrument, *P* = 6.8×10^-4^) and eGFR_Cys_ (β = 0.4% increase per 34% relative decrease in *SHROOM3* genetic instrument, *P* = 1.4×10^-4^) (Figure 2 & Supplementary Table 4). We obtained similar results in our trans-ancestry eGFR_Crea_ analysis (β = 1.4% increase per 34% relative decrease in *SHROOM3* genetic instrument, *P* = 2.7×10^-3^). Summary-level MR for eGFR were concordant with the individual-level allele score MR results in UK biobank (Supplementary Table S4 & S5). The impact of genetically predicted tubulointerstitial *SHROOM3* expression on eGFR was comparable between individuals with or without diabetes (Supplementary Table S4). Genetically predicted tubulointerstitial *SHROOM3* expression was also associated with uACR (β = 1.5% increase per 34% relative decrease in *SHROOM3* genetic instrument, *P* = 0.01) (Figure 2). We did not observe an association between genetically predicted tubulointerstitial *SHROOM3* expression and longitudinal eGFR decline (*P* = 0.47), risk of CKD (52,204 CKD cases and 884,322 controls; *P* = 0.4), or kidney volume (*P* = 0.19) (Supplementary Table S4 & S6). Phenome-wide MR demonstrated no association between genetically predicted tubulointerstitial *SHROOM3* expression and any of the 228 phenotypes following Bonferroni correction (Supplementary Table S7).

**Figure 2:**
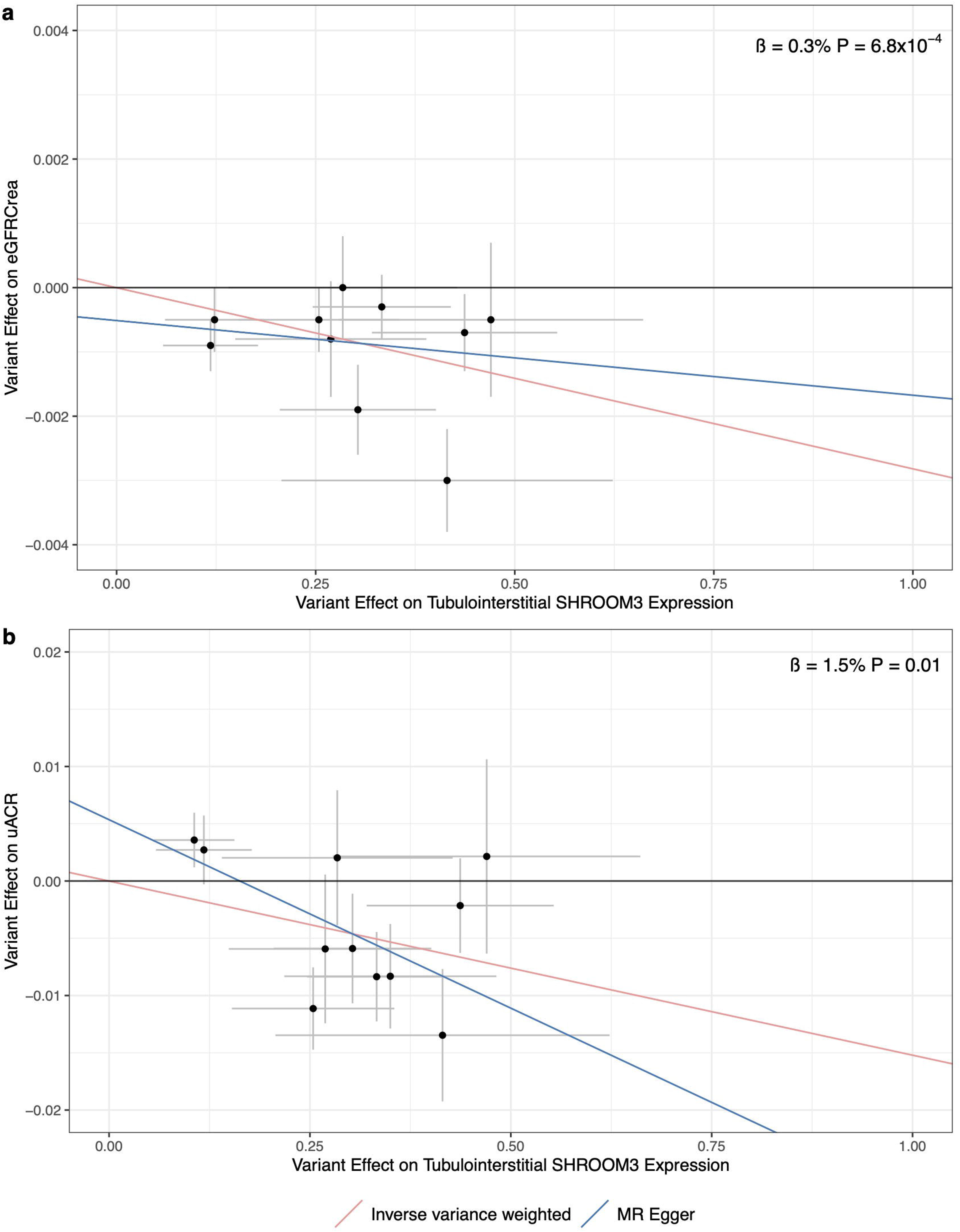
Two-sample Mendelian randomization plots of tubulointerstitial *SHROOM3* expression on (A) estimated glomerular filtration rate (eGFR_Crea_)^3^ and (B) urinary albumin-to-creatinine ratio (uACR)^5^. Each dot represents a genetic variant within the genetic instrument. X axis represents standard deviation (SD) change in *SHROOM3* expression. Y axis represents the SD change in kidney trait. Beta represents the percent change in trait per 34% relative decrease in *SHROOM3* genetic instrument score. Full results can be found in Supplementary Table S4.

### Glomerular SHROOM3 MR Analysis

MR analysis indicated a nominal association between genetically predicted glomerular *SHROOM3* expression and cross-sectional eGFR_Crea_ in the opposite direction as the tubulointerstitial instrument (β = 0.13% decrease per 34% relative decrease in *SHROOM3* genetic instrument, *P* = 0.03; *P*_heterogeneity with tubulointerstitial instrument_ < 0.001) (Figure 3 & Supplementary Table S8). In contrast, genetically predicted glomerular *SHROOM3* expression and uACR were nominally associated in the same direction as the tubulointerstitial instrument (β = 1.03% increase per 34% relative decrease in *SHROOM3* genetic instrument, *P* = 0.03, *P*_heterogeneity with tubulointerstitial instrument_ = 0.5) (Supplementary Table S8). Individual-level allele score MR from the UK biobank did not show an association between genetically predicted glomerular *SHROOM3* expression and kidney phenotypes (Supplementary Table S9). We did not observe any association between genetically predicted glomerular *SHROOM3* expression and eGFR decline, BUN, CKD, or kidney volume (all *P* > 0.05). Moreover, our phenome-wide MR (Supplementary Table S10) did not reveal any phenotypes associated with genetically predicted glomerular *SHROOM3* expression.

**Figure 3:**
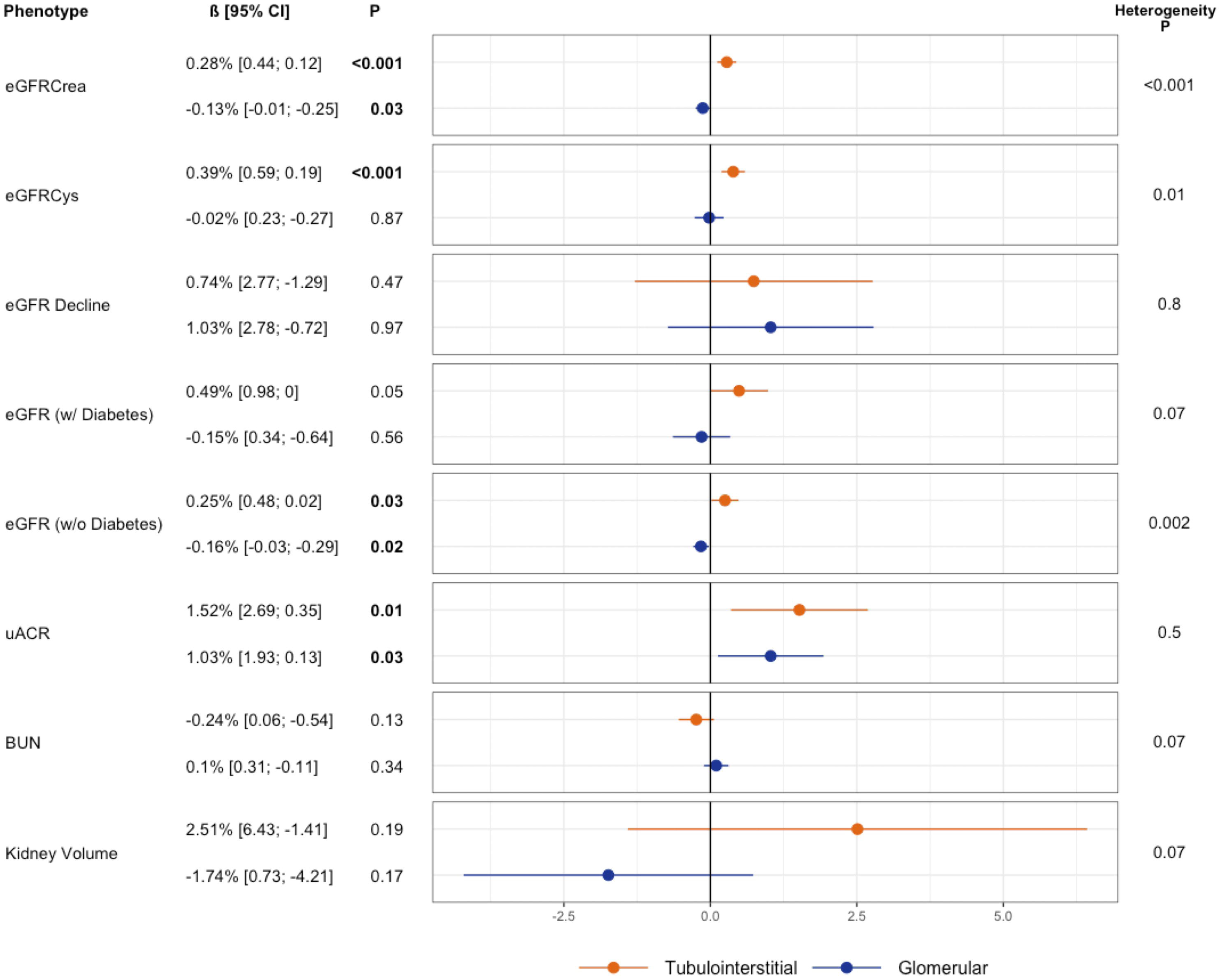
Forest plot comparing the impact of *SHROOM3* expression on kidney traits in the glomerulus versus the tubulointerstitial region reveals compartment-specific effects. Beta represents the percent change in each trait per one standard deviation decrease in the *SHROOM3* genetic instrument score. Results depicted are from the inverse variance weighted analysis. Heterogeneity P-Value determined using Cochrane’s Q statistic. eGFR_Cys_, estimated glomerular filtration rate from serum cystatin C; eGFR_Crea_, estimated glomerular filtration rate from serum creatinine; uACR, urinary albumin-to-creatinine ratio; BUN, blood urea nitrogen.

### Colocalization of SHROOM3 Genetic Variants and Kidney Outcomes

We observed strong evidence for colocalization of genetic signals for tubulointerstitial *SHROOM3* expression and eGFR_Crea_ (H_4_ = 96%), eGFR_Cys_ (H_4_ = 95%), uACR (H_4_ = 83%), and kidney volume (H_4_ = 92%; Figure 4). This colocalization suggests the presence of a shared causal variant influencing both tubulointerstitial *SHROOM3* expression and eGFR, uACR, and kidney volume. In contrast, there was no colocalization between genetically predicted glomerular *SHROOM3* expression and eGFR_Crea_ or uACR (H_4_ < 80%) (Supplementary Table S11), nor tubulointerstitial *SHROOM3* expression with glomerular *SHROOM3* expression or multi-tissue *SHROOM3* expression.

**Figure 4:**
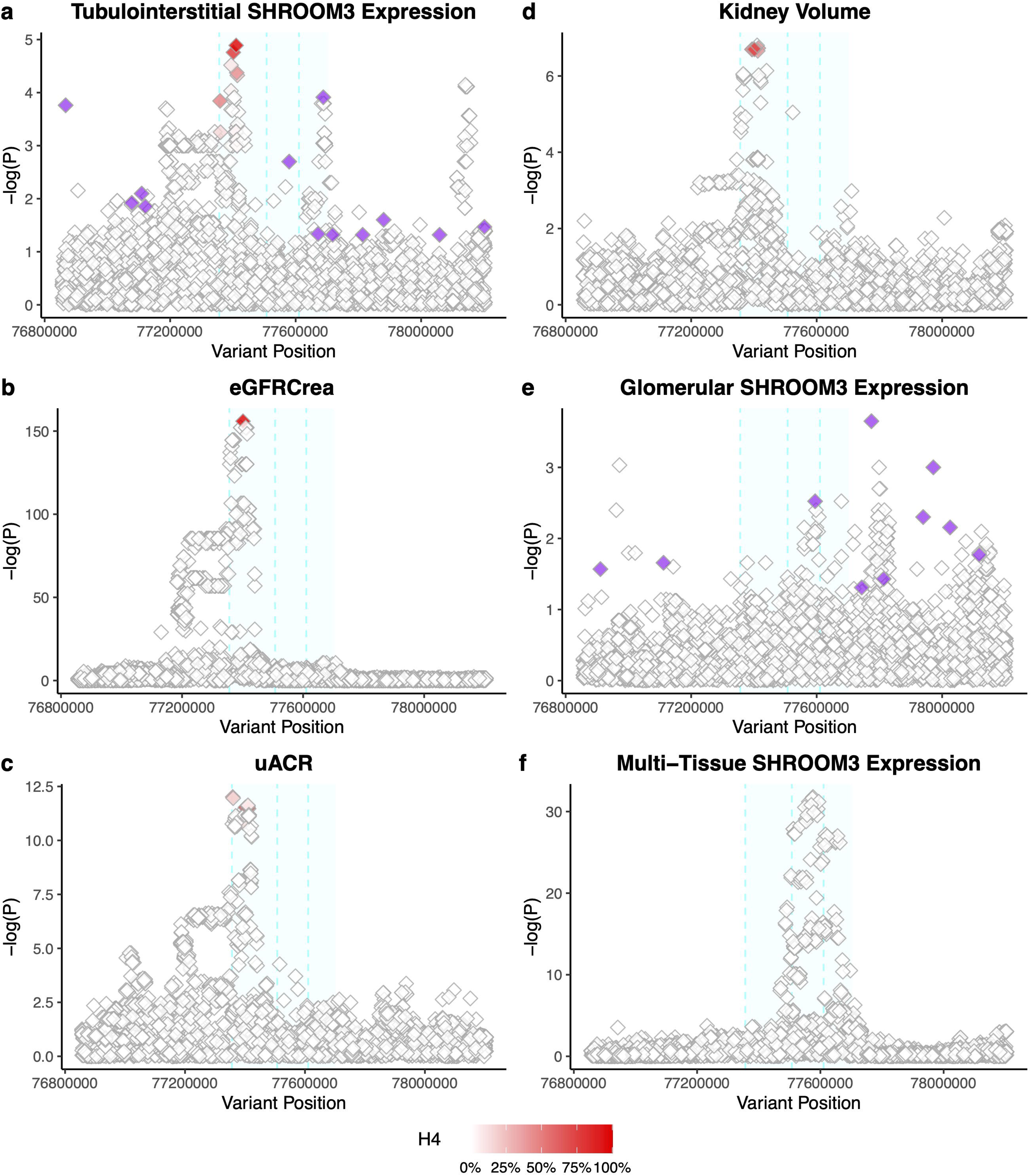
Colocalization of genetically predicted (A) tubulointerstitial *SHROOM3* expression with (B) estimated glomerular filtration rate (eGFRCrea),^3^ (C) kidney volume,^38^ (D) urinary albumin-to-creatinine ratio (uACR),^5^ (E) glomerular *SHROOM3* expression, and (F) multi-tissue *SHROOM3* expression reveals compartment-specific colocalization. Degree of redness represents the posterior probability of colocalization of the genetic variant. Purple dots represent genetic variants included in *SHROOM3* genetic instruments. The cyan shaded region represents the location of the *SHROOM3* gene on chromosome 4, while the dashed cyan lines represent the three transcription start sites within *SHROOM3* corresponding to three unique *SHROOM3* isoforms.^67^ Colocalization results can be found in Supplementary Table S11.

### SHROOM3 Rare Genetic Variant Analysis

No monogenic phenotype has been reported from rare pathogenic variants in *SHROOM3*. Homozygous *SHROOM3* loss-of-function variants were absent in the population sequencing data from gnomAD and BRAVO, including whole genome and exome data. Meta-analysis of 270,000 population sequencing participants revealed a prevalence rate of heterozygous loss-of-function *SHROOM3* variants at 0.07% (95% CI: 0.05% – 0.08%) in the general population (Supplementary Table S12), consistently across all eight available ancestries (Supplementary Table S13).

In UK Biobank exome sequencing data, we identified 3435 individuals carrying a rare loss-of-function or predicted pathogenic missense variant in *SHROOM3*. These variants were nominally associated with eGFR_Cys_ (β = 3.44% increase in trait in *SHROOM3* rare variant carriers, *P* = 0.03) but not with uACR (*P* = 0.26) (Table 1).

**Table 1:**
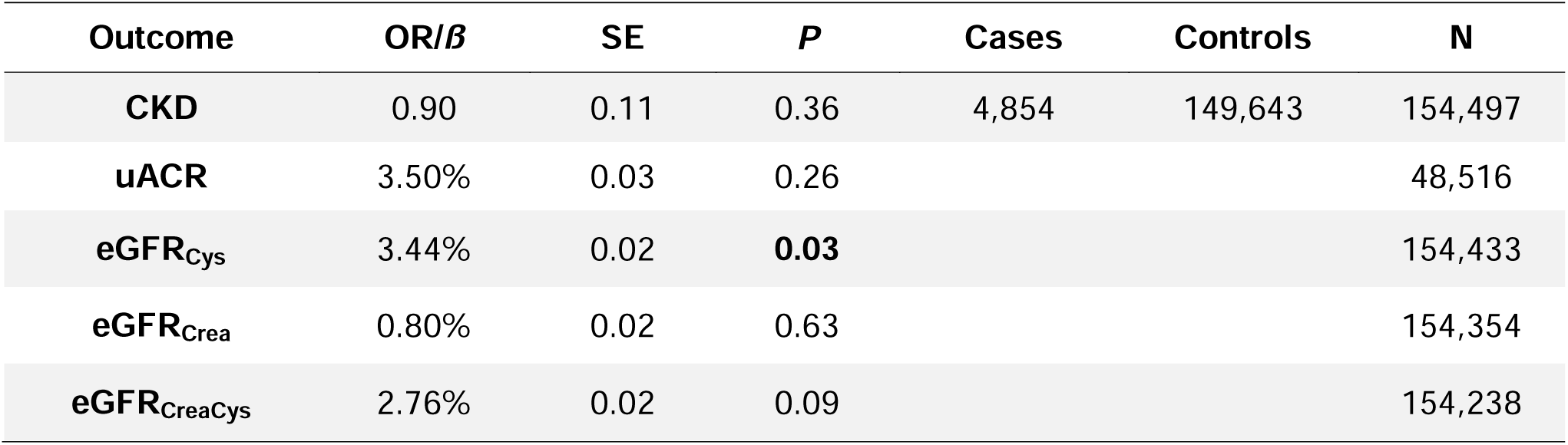
Rare variant association analysis of *SHROOM3* pathogenic variants shows weak association with estimated glomerular filtration rate using serum cystatin C (eGFR_Cys_). Rare variants were classified as predicted loss-of-function (nonsense, frameshift, and splice donor or acceptor) variants or missense variants with an MCAP score of > 0.025 indicating pathogenicity. Beta (*ß*) represents % change in trait if carrying a *SHROOM3* rare variant. N represents the number of samples included in the analysis. 3,435 rare pathogenic variants were present in the UK Biobank exome sequencing database. N, sample size; CKD, chronic kidney disease; uACR, urinary albumin-to-creatinine ratio eGFR_Crea_, estimated glomerular filtration rate from serum creatinine; eGFR_CreaCys_, estimated glomerular filtration rate from serum cystatin C and creatinine.

### Tubulointerstitial SHROOM3 Impacts on the Circulating Proteome

In a novel analysis looking at the impact of genetically predicted *SHROOM3* expression on the circulating proteome, the three most significant associations with genetically predicted tubulointerstitial *SHROOM3* expression were with proteins encoded by genes located on chromosome 4 and within 500 kilobases of *SHROOM3* (*NAAA*, *SCARB2*, and *ART3*) (Table 2 & Supplementary Table S14). Reduced collagen type XVIII alpha one (COL18A1), a protein biomarker located on chromosome 21, was also associated with lower genetically predicted tubulointerstitial *SHROOM3* expression and surpassed Bonferroni-adjusted significance (β = -2.15% per 34% relative decrease in *SHROOM3* genetic instrument, *P* = 1.2×10^-5^). Genetically predicted glomerular *SHROOM3* expression was not associated with changes in a circulating biomarker (Supplementary Table S15).

**Table 2:**
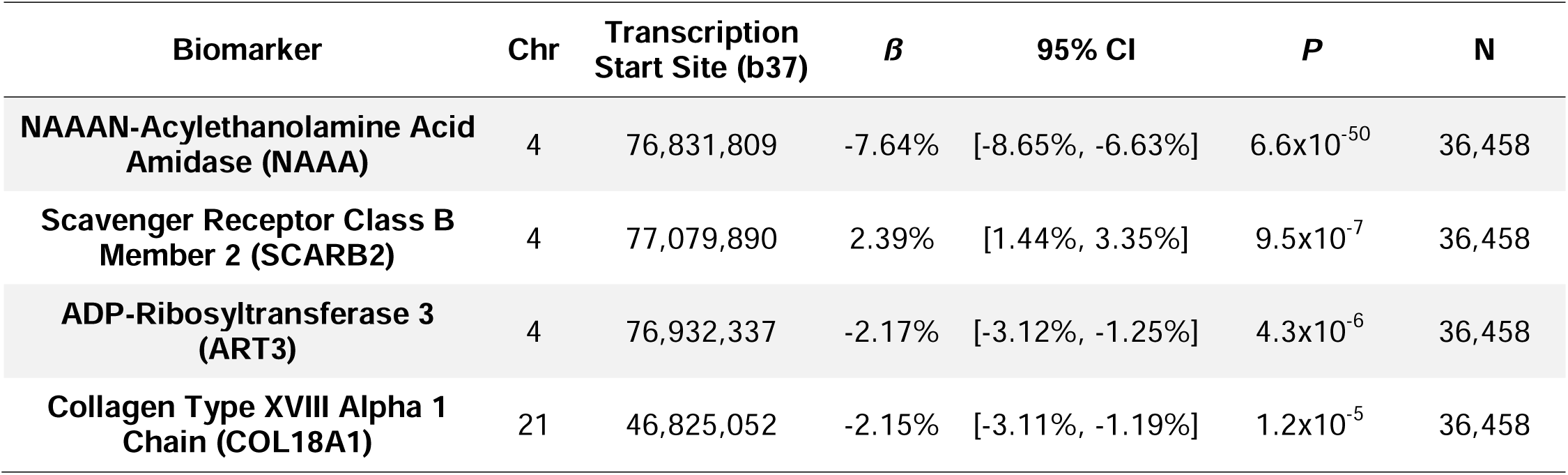
The impact of genetically predicted tubulointerstitial *SHROOM3* expression on proteome-wide serum biomarkers in European ancestry UK Biobank participants. Beta (*ß*) represents percent change in biomarker per 34% relative decrease in *SHROOM3* genetic instrument score. N represents sample size. Chr represents chromosome of gene encoding biomarker. *NAAA*, *SCARB2*, and *ART3* are genes located adjacent to the *SHROOM3* gene (chr4:77,356,253-77,704,406). Only biomarkers surpassing the Bonferroni-corrected threshold of *P* < 3×10^-5^ are shown. Proteome-wide results can be found in Supplementary Table S14.

### Immunofluorescence Staining Validation of COL18A1 Association

The association of *SHROOM3* expression with COL18A1 was reinforced experimentally by visualizing the expression pattern of Col18a1 in the kidneys of 3-month-old *Shroom3^Gt/+^* knockout mice. Wild type mice kidneys demonstrated robust Col18a1 protein expression surrounding the parietal epithelium of the renal corpuscle (glomerulus and Bowman’s capsule) and around the tubular epithelium. *Shroom3^Gt/+^*knockout kidneys exhibited a similar, but reduced, pattern of Col18a1 protein expression around the renal corpuscle. Notably, Col18a1 expression was even more reduced around the cortical and medullary tubular epithelium (Figure 5). The staining patterns indicate a reduction in Col18a1 protein levels in the kidneys resulting from *Shroom3* heterozygous knockout.

**Figure 5:**
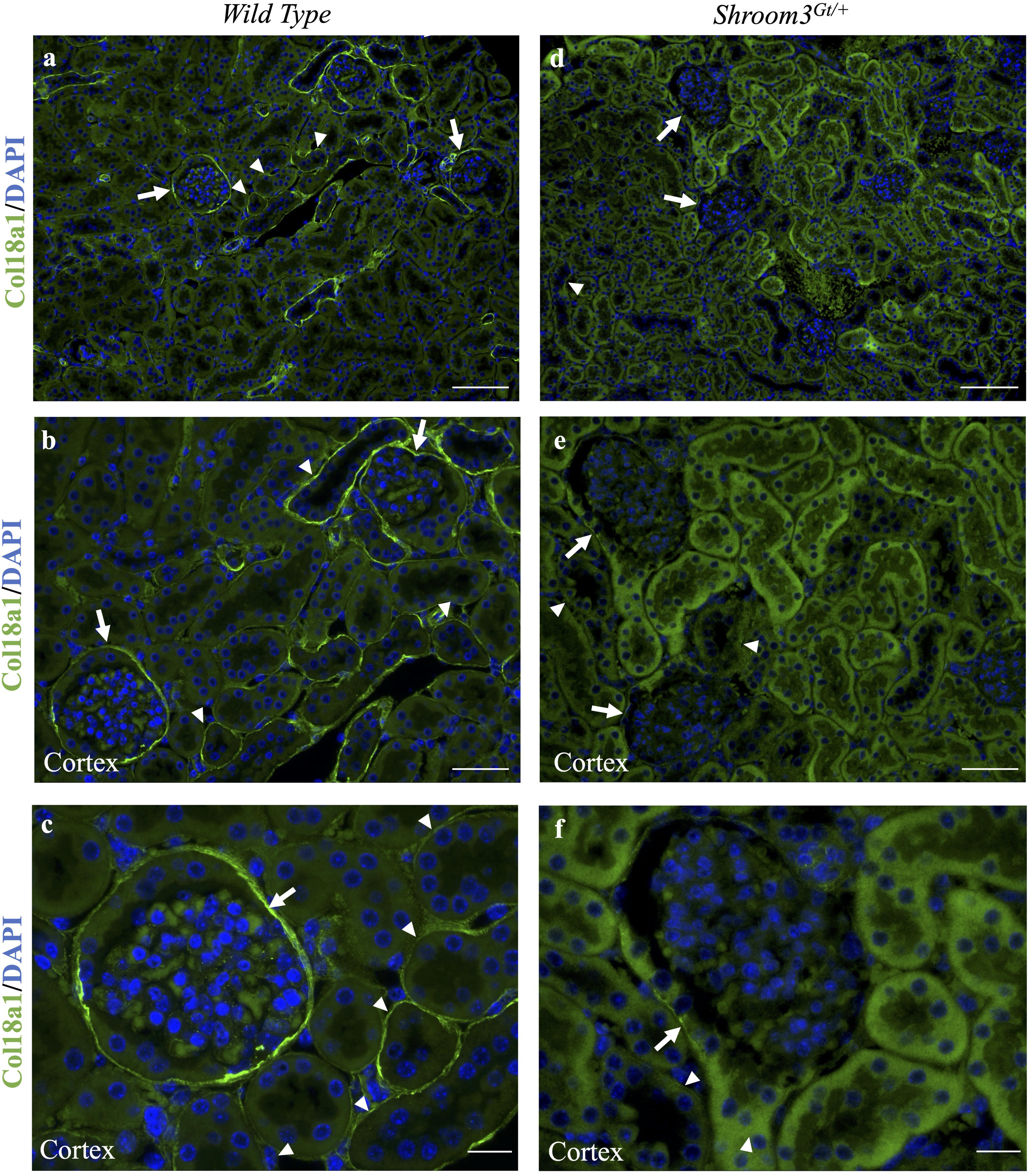
Immunofluorescence staining of Col18a1 protein in *Shroom3* heterozygous knockout mice shows reduced Col18a1 protein expression around the kidney glomeruli and tubules. Col18a1 immunofluorescence staining was performed in adult 3-month-old wild type and *Shroom3* heterozygous knockout (*Shroom3^Gt/+^*) kidneys (n=2). Fluorescent green staining represents Col18a1 protein expression. **(A-C)** In wild type kidneys, Col18a1 protein expression prominently surrounds the renal corpuscle (glomerulus and Bowman’s capsule) (arrow) and tubules (arrowhead) in the cortical and medullary kidney regions. (D-F) *Shroom3^Gt/+^* kidneys show substantially reduced expression of Col18a1 around the glomeruli and even less expression around the tubules. Scale bars = 100 µm (A, D). Scale bars = 50 µm (B, E). Scale bars = 20 µm (C, F).

## Discussion

Variants that impact *SHROOM3* expression in the kidney tubulointerstitial and glomerular regions do not colocalize, suggesting different genetic regulatory mechanisms. Leveraging this distinction, we were able to evaluate genetic expression specific to these compartments by creating separate genetic instruments for targeted spatial evaluation in multi-omic MR analyses. We observed a 34% relative reduction in genetically predicted tubulointerstitial *SHROOM3* expression led to a 0.28% increase in cross-sectional eGFR, 1.52% increase in uACR, and no detectable association with eGFR decline nor prevalent CKD. Colocalization analysis supported genetically predicted tubulointerstitial expression, rather than glomerular, drove the regional GWAS associations. Evaluating 1,463 biomarkers in the human circulating proteome, lower genetically predicted *SHROOM3* expression led to less COL18A1 in the blood. We then validated this causal relationship by immunofluorescence staining the kidneys of *Shroom3* knockout mice, confirming the link and temporal sequence of reduced Col18a1 from reduced *Shroom3* expression.

Cross-sectional eGFR is determined by a combination of two factors: maximally attained eGFR, determined by nephron endowment, and the rate of eGFR decline due to age or disease.^49^ Gorski *et al*.’s 2022 study categorized variants associated with eGFR into two groups: 1) those associated with both maximally attained eGFR and its subsequent decline, and 2) those associated with maximally attained eGFR but not with eGFR decline.^36^ Three genes, *GATM*, *CPS1*, and *SHROOM3* fell into the second category, highlighting a potential role in affecting nephron endowment. Building on these findings, we found reduced genetically predicted *SHROOM3* expression in the tubulointerstitial is associated with increased uACR and increased cross-sectional eGFR rather than eGFR decline. The question arises: does *SHROOM3* influence nephron endowment and, if so, what is the result?

Prior research identified *SHROOM3* as a key factor in kidney development. Human embryonic kidneys show the highest *SHROOM3* expression in the nephron progenitor cells, which give rise to all segments of the nephron.^50,51^ Both *Shroom3* heterozygous and homozygous knockout mice demonstrate significant dose-dependent reductions in glomerular number compared to wild type mice at embryonic day 18.5, with many glomeruli becoming cystic or collapsing.^9^ Although the exact mechanism is unknown, it is possible that the reduction of *SHROOM3* in the nephron progenitor cells results in the improper development of nephrons. We also demonstrated an association between *SHROOM3* and COL18A1, an extracellular collagen protein with a known role in kidney development. In embryonic mice, the absence of Col18a1 leads to altered kidney development, including a shorter ureteric tree, smaller kidney volume, reduced nephron count, and fewer terminal branch points and ureteric tips.^48^ Although MR analysis did not reveal a significant association between tubulointerstitial *SHROOM3* expression and total kidney volume, genetic colocalization of the phenotypes suggests the possibility of a mediating factor, possibly COL18A1. The connection between Shroom3 and Col18a1 in the tubulointerstitial area of postnatal mice suggests changes in Shroom3 expression could impact nephron formation via Col18a1. One possible hypothesis is disruptions in the ability of developing and mature nephrons to connect to the extracellular matrix. The observed interactions between *SHROOM3* and COL18A1 warrant further investigation to understand the pathways that influence kidney morphology.

COL18A1 and its cleavage product, endostatin, an antiangiogenic protein derived from the C-terminal of COL18A1, have significant roles in kidney disease pathology and function.^52^ In young mice with induced glomerulonephritis, *Col18a1* knockout led to heightened inflammatory responses and a decrease in glomerular and interstitial vascular density.^53^ A study of 7-month-old *Col18a1* knockout mice demonstrated podocyte foot process effacement and loosening of the proximal tubular basement membrane.^54^ In adult diabetic mice, administering endostatin peptide resulted in significant reductions in glomerular hypertrophy, hyperfiltration, and albuminuria.^55^ Although *SHROOM3* and COL18A1 have imperfectly understood individual roles in CKD, the interplay between both in CKD pathogenesis, embryonically and post-natally, warrants further exploration.

Altered nephron endowment exerts a distinctive impact on kidney phenotypes. According to the Brenner hypothesis, a diminished nephron count leads to a reduction in the total filtration surface area, prompting an adaptive increase in single nephron glomerular filtration rate (snGFR).^56,57^ This adaptive response leads to ‘relative’ hyperfiltration to normalize the overall eGFR despite a reduced number of functioning nephrons.^58^ The increase in snGFR is relative to the amount of renal mass that has been lost. Over years or decades, an increased snGFR causes sclerosis in the glomeruli of the remaining nephrons, proteinuria, hypertension, and CKD. In contrast to CKD, where low eGFR is associated with higher ACR, hyperfiltration manifests initially with increased eGFR and elevated uACR.^58,59^ This pattern of increased eGFR and uACR is also observed in early stages of diabetic nephropathy and focal segmental glomerulosclerosis.^59,60^ Our findings support hyperfiltration, with reduced tubulointerstitial *SHROOM3* expression associated with both increased uACR and cross-sectional eGFR, likely stemming from a reduction in nephron mass during kidney development (Figure 6).

**Figure 6:**
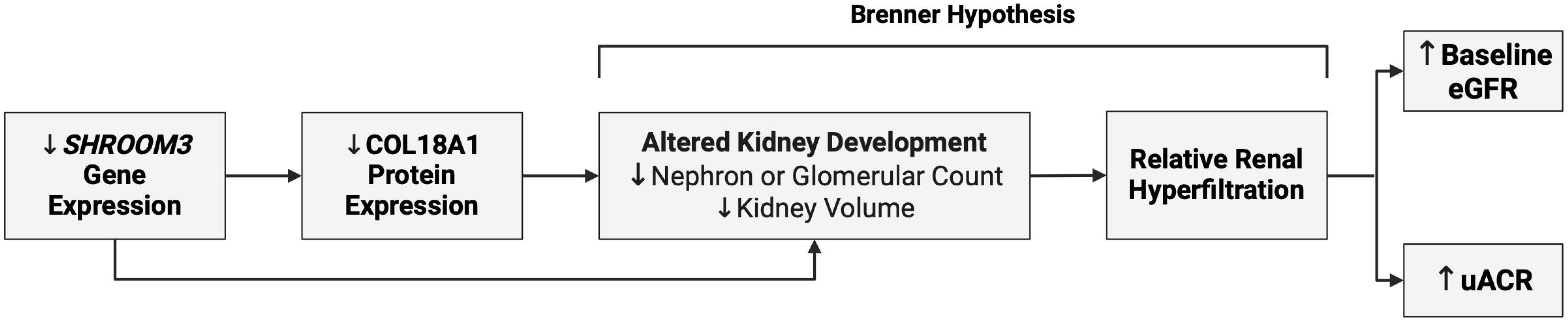
Proposed pathway depicting the impact of *SHROOM3* gene expression on estimated glomerular filtration rate (eGFR) and urinary albumin-to-creatinine ratio (uACR) through kidney development and hyperfiltration. The association of *Shroom3* expression with altered kidney development was previously demonstrated in Khalili et al., 2016,^9^ while the association of Col18a1 expression with altered kidney development was shown in Rinta-Jaskari et al., 2023.^48^ Created with BioRender.com.

The absence of homozygous loss-of-function *SHROOM3* variants in population sequencing of 270,000 human participants is consistent with neonatal lethality observed in *Shroom3* homozygous knockout mice.^45^ Due to the lethality of homozygous knockout, our rare variant analysis was limited to individuals with heterozygous variants. Rather than examining the quantity of *SHROOM3,* rare exonic missense variants in *SHROOM3* would be expected to impact protein function and were nominally associated with higher cross-sectional eGFR, but were not found to be associated with prevalent CKD in the healthy UK Biobank population.

Despite widespread expression of *SHROOM3* RNA and protein across human tissues, our phenome-wide MR did not uncover significant associations between altered genetically predicted *SHROOM3* expression and any of the 228 examined dichotomous outcomes. The lack of findings may be attributable to the tissue specificity of the *SHROOM3* genetic instruments. Complex diseases are more susceptible to the influence of tissue-specific effects, particularly those tissues relevant to the disease.^61^ Our kidney compartment-specific genetic instruments are more potent in assessing relationships with kidney diseases compared to other traits. While a multi-tissue genetic instrument has more power and might theoretically represent gene expression in all tissues, in reality, serious heterogeneity arises from varying eQTL effects across tissues.^62,63^ Furthermore, recent expansions in human kidney single cell RNA sequencing databases have allowed for the development of sufficiently powered kidney compartment-specific genetic instruments.^4,24^

Strengths of this work include large non-overlapping population-based cohorts, utilization of tissue compartment-specific eQTL genetic instruments for spatial multi-omics, integration of colocalization, phenome-wide, proteome-wide and rare variant analyses of *SHROOM3* and validation of findings in a *Shroom3* knockout mouse. There is an increasing recognition of the pivotal role of tissue-specific and cell-type-specific eQTLs in gene regulation, which are more likely to colocalize with and explain GWAS signals.^64-66^ Recently, tissue-specific analyses identified kidney tubulointerstitial *DAB2* expression as a causal risk factor for CKD, a connection not previously recognized when analyzing whole kidney expression.^66^ Utilizing MR offers an advantage to GWAS, as it employs multiple variants to examine the correlation between the effects of these variants on gene expression and their influence on various outcomes rather than evaluating each variant independently as in GWAS yielding a theoretical improvement in power. Furthermore, MR is inherently robust against reverse causation and confounding factors, a notable strength compared to conventional epidemiological methods.^13^ Limitations of this study include reliance on largely European ancestry GWAS data, though we did validate our findings with eGFR_Crea_ in a trans-ancestry GWAS, and the constrained sample size for the kidney compartment eQTL data.

In conclusion, human population and knockout mice studies support a novel interaction between SHROOM3 and COL18A1 leading to impaired kidney development and hyperfiltration.

## Data Availability

CKD Genetics Consortium summary-level data is publicly available (https://ckdgen.imbi.uni-freiburg.de/) and UK Biobank individual-level data is available with application (https://www.ukbiobank.ac.uk/enable-your-research/apply-for-access). UK Biobank phenome-wide summary statistics are available (https://www.leelabsg.org/resources). eQTL summary-level data obtained from NephQTL2 and GTEx is also publicly available (http://nephqtl2.org) (https://gtexportal.org/home/).

## Acknowledgements

We would like to thank Dr. Thomas Drysdale at the University of Western Ontario for gifting the heterozygous *Shroom3* knockout mice and Dr. Satu Kuure at the University of Helsinki and Dr. Takako Sasaki at Oita University for gifting the Col18a1 antibody. The authors thank the participants and investigators of the contributing consortiums, including NEPTUNE, UK Biobank, the CKD Genetics Consortium, and GTEx. The GTEx Project was supported by the Common Fund of the Office of the Director of the National Institutes of Health, and by NCI, NHGRI, NHLBI, NIDA, NIMH, and NINDS. The data used for the analyses described in this manuscript were obtained from: the GTEx Portal on 11/11/2022.

## Author Contributions

MBL, DB, and PSG designed the project. AP and DB performed the mice experiments and imaging. PSG performed all statistical analyses and drafted the manuscript. MBL revised the manuscript. All authors provided intellectual content and approved the final version of the manuscript.

## List of Supplementary Materials

**Supplementary Table S1.** STrengthening the Reporting of OBservational studies in Epidemiology using Mendelian Randomization (STROBE-MR) Checklist.

**Supplementary Table S2.** Genetic instrument for tubulointerstitial SHROOM3 expression and glomerular SHROOM3 expression.

**Supplementary Table S3.** Phenoscanner V2 search of tubulointerstitial and glomerular genetic instrument variants.

**Supplementary Table S4.** Summary-level Mendelian randomization results using tubulointerstitial SHROOM3 genetic instrument.

**Supplementary Table S5.** Individual-level allele score Mendelian randomization using UK Biobank phenotype data and tubulointerstitial genetic instrument.

**Supplementary Table S6.** Inverse variance weighted meta-analysis of CKD Mendelian randomization results across CKD Genetics Consortium and UK Biobank.

**Supplementary Table S7.** Phenome-wide Mendelian randomization of 228 binary phenotypes in the UK Biobank using the tubulointerstitial genetic instrument.

**Supplementary Table S8.** Summary-level Mendelian randomization results using glomerular SHROOM3 genetic instrument.

**Supplementary Table S9.** Individual-level allele score Mendelian randomization using UK Biobank phenotype data and glomerular genetic instrument.

**Supplementary Table S10.** Phenome-wide Mendelian randomization of 228 binary phenotypes in the UK Biobank using the glomerular genetic instrument.

**Supplementary Table S11.** Colocalization of tubulointerstitial and glomerular genetic instruments with kidney phenotypes.

**Supplementary Table S12.** Prevalence of predicted loss-of-function SHROOM3 variants in the general population.

**Supplementary Table S13.** Prevalence of predicted loss-of-function SHROOM3 variants across eight ancestries present in gnomAD v2.1.

**Supplementary Table S14.** Proteome-wide individual-level allele score Mendelian randomization using UK Biobank biomarker data and tubulointerstitial genetic instrument.

**Supplementary Table S15.** Proteome-wide individual-level allele score Mendelian randomization using UK Biobank biomarker data and glomerular genetic instrument.

